# Data Matters: The Impact of Data Curation in the Classification of Histopathological Datasets

**DOI:** 10.64898/2026.04.16.26351016

**Authors:** Daniel Brito-Pacheco, Panos Giannopoulos, Constantino Carlos Reyes-Aldasoro

**Affiliations:** School of Science and Technology, City St. George’s, University of London, EC1V 0HB, London, United Kingdom; The Institute of Cancer Research, Integrated Pathology Unit, Division of Molecular Pathology, Sutton, United Kingdom

## Abstract

In this work, the impact of outliers on the performance of machine learning and deep learning models is investigated, specifically for the case of histopathological images of colorectal cancer stained with Haematoxylin and Eosin. The evaluation of the impact is done through the systematic comparison of one machine learning model (Random Forests) and one deep learning model (ResNet-18). Both models were trained with the popular NCT-CRC-HE-VAL-100K dataset and tested on the CRC-HE-VAL-7K companion set. Then, a curation process was performed by analysing the divergence of patches based on chromatic, textural and topological features of the training set and removing outliers to repeat the training with a cleaned dataset. The results showed that machine learning models, can benefit more from improvements in the quality of data, than deep learning models. Further, the results suggest that deep learning models are more robust to outliers as, through the training process, the architectures can learn features other than those previously mentioned.

## 1 Introduction

The success of machine learning (ML) and deep learning (DL) in the computational analysis of pathology images has relied heavily on the availability of large, annotated datasets (e.g., [2, 7, 14, 18, 20, 23]). It is because of this that robust and generalisable models that are able to classify images stained in this way are particularly important. For this purpose, openly-available datasets are of great value to the computational pathology community. Among these, the NCT-CRC-HE-100K dataset and its companion test set, CRC-VAL-HE-7K [10, 11], have been widely used (more than 33,000 downloads at the time of writing) as benchmarks for the classification of colorectal cancer histology images [12, 19, 22]. These datasets have enabled the development and comparison of a wide range of models, particularly convolutional neural network (CNNs).

In this context, prior work has highlighted potential limitations of the aforementioned datasets (NCT-CRC-HE-100K and CRC-VAL-HE-7K) [6, 9]. In particular, differences in data quality and preprocessing, led to differences in class separability between the training and testing sets. In those studies, the datasets were analysed using colour, histograms, topological and texture-based features, revealing inconsistencies in the distribution of classes between the two sets among other problems like severe JPEG artifacts and corrupted tissue samples resulting from incorrect image dynamic range handling. Specifically, the 7K-set exhibited a higher degree of class separability than the 100K-set, suggesting that the datasets may not be directly comparable and that models trained on one may not generalise well to the other.

These previous findings raise important questions: *to what extent do the outliers, and differences in class separability affect the performance of ML and DL models? In particular, does the presence of samples which may be atypical due to issues such as mislabelling or poor normalisation in certain classes, affect the generalisation of ML models to other datasets?*

In this paper, the impact of dataset curation is studied through classification performance of models trained on altered variations of the training set. Using a combination of hand-crafted features, derived from Persistent Homology (PH), Gabor filtering, and colour histograms, outliers were first identified within specific tissue classes based on statistical deviations in feature space. Then, the performance of a classical ML model (Random Forest classifier), was used to evaluate the effect of removing these outlier samples on ML models. Furthermore, to assess whether these effects extend to DL approaches, similar experiments were performed using a ResNet-18 architecture. Since all models are trained on one dataset (100K-set), then shown a completely new dataset (7K-set), the accuracy on the test set is used as a score for generalisation. By comparing models trained on the original and cleaned datasets, the results revealed that the traditional ML model can potentially benefit more from a clean dataset, than the DL model, when it comes to generalisation.

## 2 Materials and Methods

### 2.1 Materials

Two sets of images from colorectal cancer patients were used. Each set contains patches of 224 *×* 224 pixels of tissues separated into nine classes: ADI (adipose tissue), BACK (background), DEB (debris), LYM (lymphocytes), MUC (mucus), MUS (smooth muscle), NORM (normal colon mucosa), STR (cancer-associated stroma), TUM (colorectal adenocarcinoma epithelium). The first set, called NCT-CRC-HE-100K, contains 100,000 patches and was sourced from 86 H&E slides the NCT biobank and the UMM pathology archive. The second set, called CRC-VAL-HE-7K, contains 7,180 patches and was sourced from 25 H&E slides from the DACHS study in the NCT biobank. These datasets are available on Zenodo [10]. Representative patches of both sets are shown in Fig. 1. Henceforth, they will be simply referred to as 100K-set and 7K-set, respectively.

**Fig. 1.**
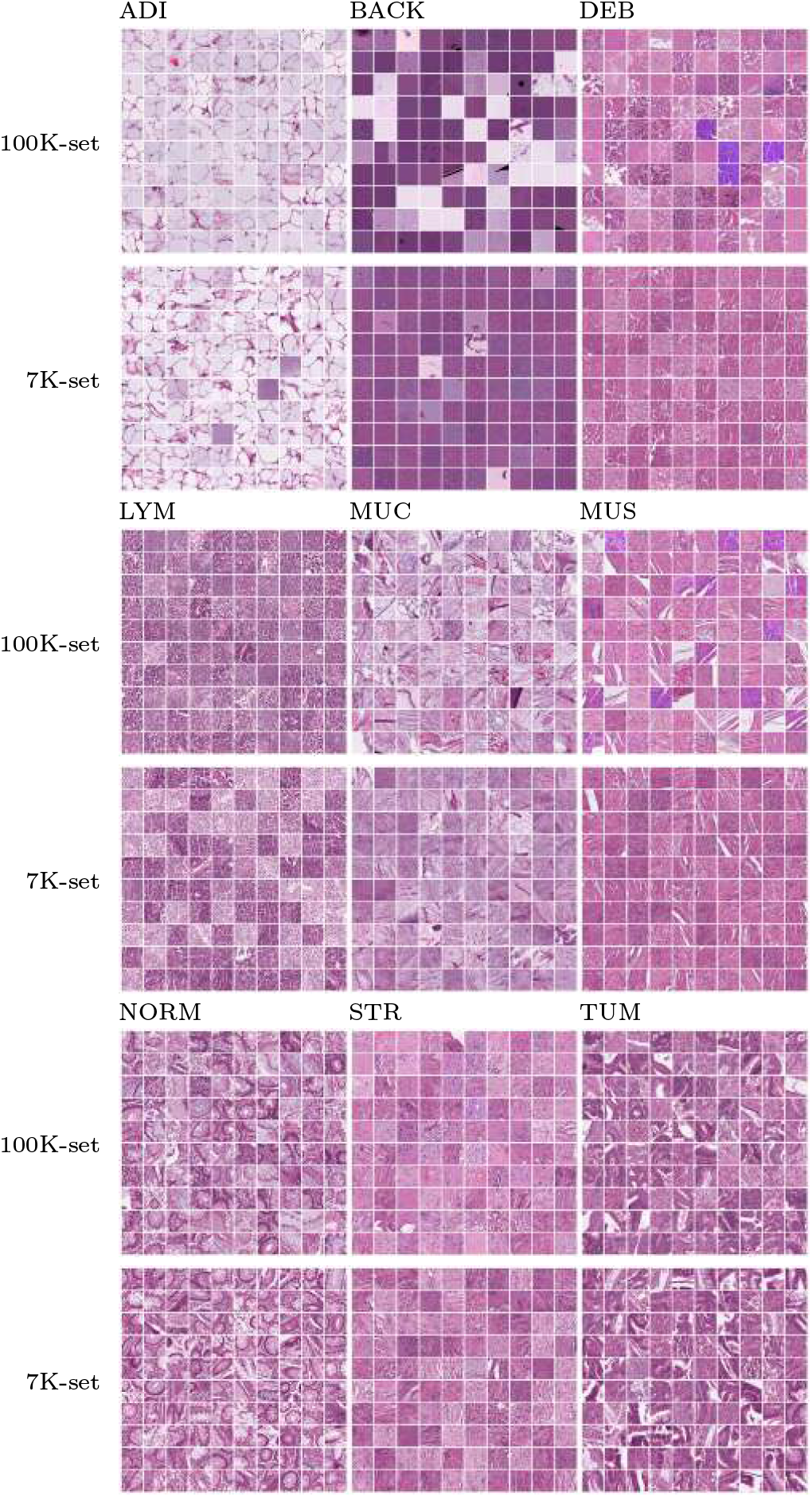
Illustration of the datasets with 100 sample patches per class from each set. By class: NCT-CRC-HE-100K images are shown above, normalized CRC-VAL-HE-7K images are shown below. ADI: adipose tissue; BACK: background; CRC: colorectal cancer; DEB: debris; LYM: lymphocytes; MUC: mucus; MUS: smooth muscle; NORM: normal colon mucosa; STR: cancer-associated stroma; TUM: colorectal adenocarcinoma epithelium.

In [11], where the details of the datasets were first published, the 100K-set was explicitly reported to have been normalised using Macenko’s method. Using the same reference image as the 100K-set, the 7K-set was subsequently normalised using the same method on a per-patch basis. For all the experiments in this study, the original 100K-set as provided on Zenodo, and the normalised 7K-set were used.

### 2.2 Methods

#### HSV Decomposition

The first set of features extracted from each patch was derived from its colour information. Each RGB patch was converted to HSV colour space, and a 16-bin histogram was computed for each channel (Fig. 2). The pixel frequencies were normalised, and the three normalised histograms were concatenated to form a 48-dimensional vector.

**Fig. 2.**
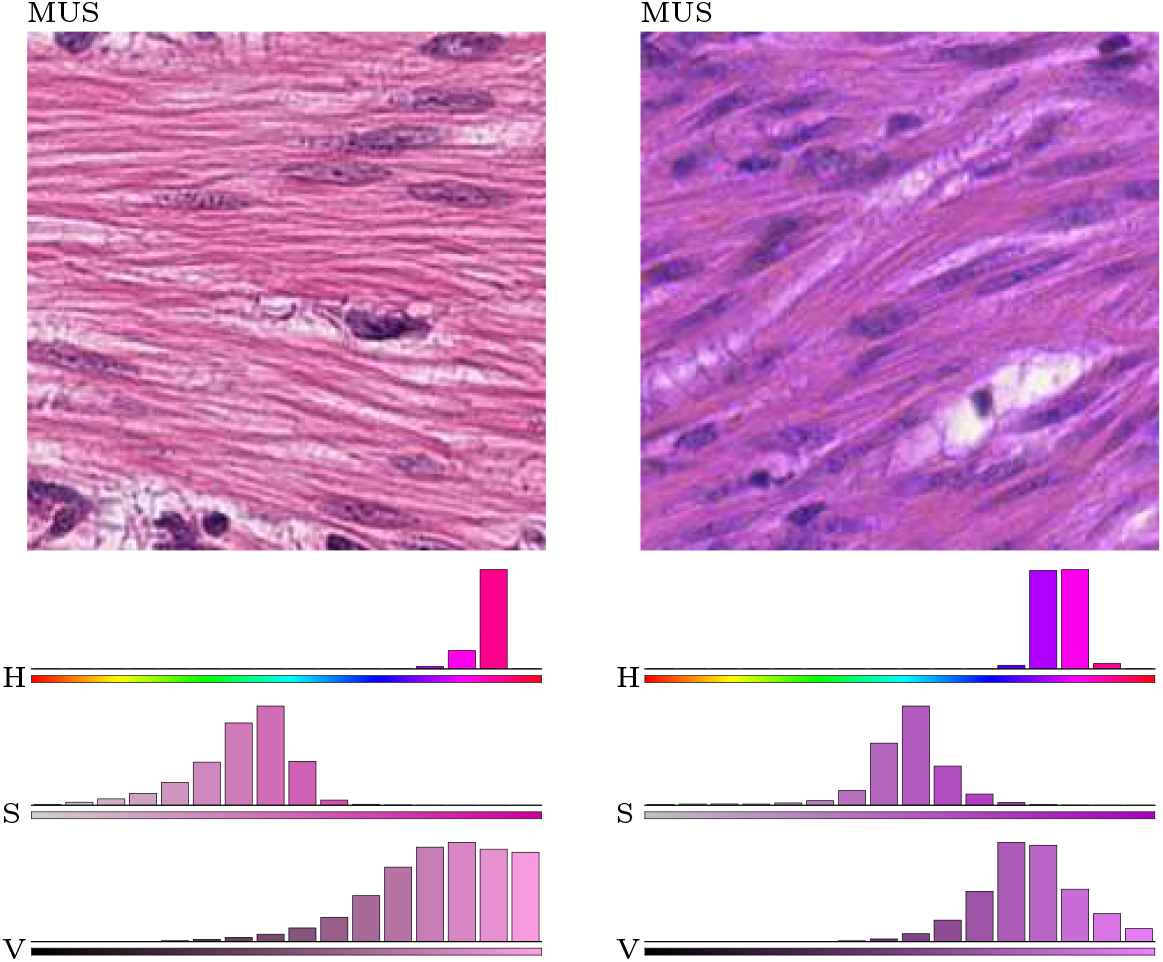
Illustration of the extraction of colour features from histological images. The histograms of each of the HSV (H: Hue, S: Saturation, V: Value) channels were calculated for two patches of the same class (MUS) with different visual characteristics. All histograms were composed of 16 bins. Whilst the two patches belong to the same class, the histograms are quite distinct.

#### Persistent Homology (PH)

PH is a mathematical tool developed in the field of Topological Data Analysis. In the context of image analysis, it allows for the extraction of topological features from images. In this subsection, a brief overview of the features extracted from the 100K-set and 7K-set is presented. For a deeper introduction to the feature extraction process using PH, the reader is referred to [6] or [15].

For each patch, PH features were extracted by constructing what is known as a persistence diagram, which captures how the topology of the image changes as a threshold is applied at different intensities of the images. Using the toy example in Fig. 3, the construction of said diagram is explained as follows. Each patch was first converted to grayscale, then a 5 × 5 median filter was applied to reduce noise and make regions of similar intensities smoother. From each smoothed image, *I* (Fig. 3 (a)), a sequence of binary images (*I*_*k*_) was obtained by applying an upper threshold on the image through 256 intensity levels (Fig. 3 (c),(d)). Each *I*_*k*_ was then converted to a topological space *T*_*k*_ by the simple rules: (1) place a vertex at every white pixel, (2) place an edge between two neighbouring pixels (diagonals are valid), (3) place a triangle when a closed cycle of three edges is present (Fig. 3 (e)). The sequence of topological spaces (*T*_*k*_), in mathematical vocabulary is known as a *filtration* (Fig. 3 (f)).

**Fig. 3.**
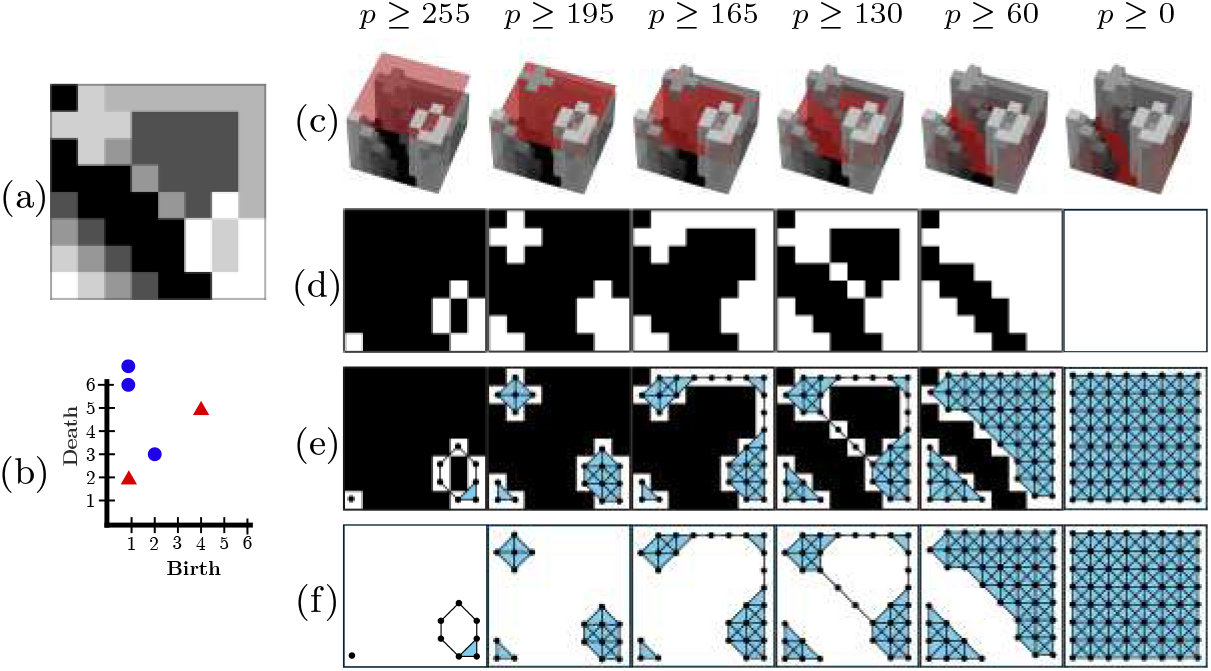
Illustration of the filtration and PH calculation. (a) Original greyscale image in the range [0, 255]. (b) Persistence diagram. (c) 3D representation of the greyscale and a threshold. (d) Binary images of pixels above the threshold. (e) Filtration overlaid on the binary images. A vertex is placed at each white pixel. Edges are added between neighbouring pixels. A triangle is added when cycles of three edges are formed. (f) Filtration with pixels removed.

The persistence diagram (Fig. 3 (b)) is a scatter plot constructed by tracking the change of topological objects (components and holes) in the filtration. When a component or hole appears for the first time, it is said to be “born”. When a component joins another, the youngest one is said to “die”. Similarly, when a hole is filled in, it is also said to die. A point is placed in the persistence diagram for every component and hole, with the coordinates of its birth on the horizontal axis, and its death on the vertical axis. In this article, blue circles were used for points corresponding to components and red triangles were used for points corresponding to holes in the persistence diagram. In Fig. 3 (f), a hole is born in the bottom right corner of the first step, which in the next step it has died (it was filled in), therefore a red triangle is placed with coordinates (1,2).

Similarly, the component in the top left corner born in step two, joins the older component in the bottom right corner in the next step, so a blue circle is placed with coordinates (2,3). Finally, in every persistence diagram, there is exactly one blue circle with death at *∞*, this represents the last component alive in the filtration, but for the computations done in this study, this point was discarded. In practice, the persistence diagram was computed using the GUDHI package for Python [16]. Examples of persistence diagrams computed for different classes are shown in Fig. 4.

**Fig. 4.**
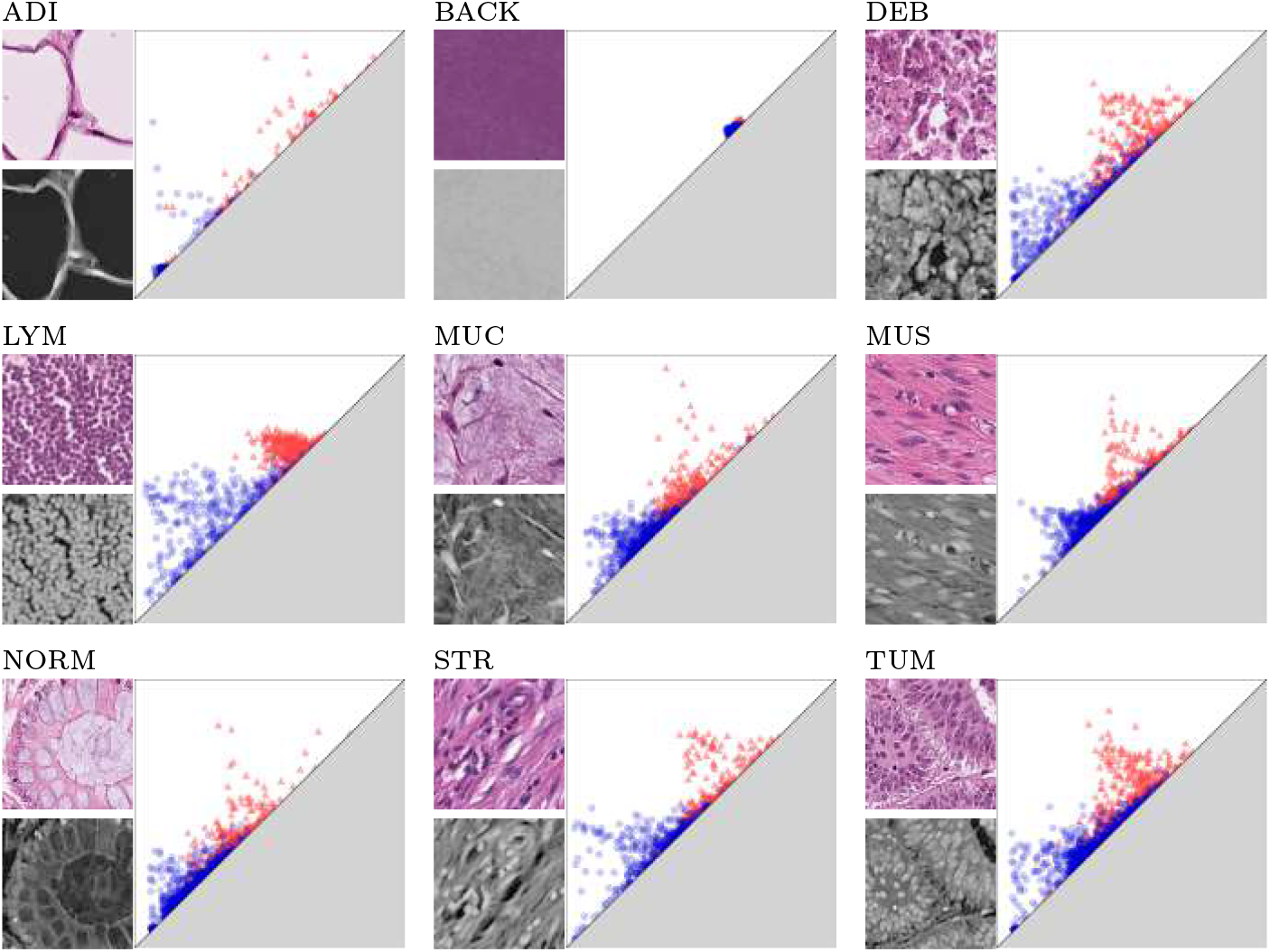
Illustration of the persistence diagrams of the histological tissues from the 100K-set. One representative patch from each of the classes (ADI, BACK, DEB, LYM, MUC, MUS, NORM, STR, TUM) is converted to greyscale and inverted. Noise is removed on the greyscale image by applying a 5 × 5 median filter. A persistence diagram is calculated from the smoothed greyscale image where blue circles are components and red triangles are holes. The distribution of the scatterplots in the persistence diagram capture differences in the textures of different tissues. ADI: adipose tissue; BACK: background; CRC: colorectal cancer; DEB: debris; LYM: lymphocytes; MUC: mucus; MUS: smooth muscle; NORM: normal colon mucosa; STR: cancer-associated stroma; TUM: colorectal adenocarcinoma epithelium.

For each patch, statistical features from its corresponding persistence diagram were computed. These were: number of components/holes, mean birth of components/holes, mean death of components/holes, standard deviation of the births of components/holes, standard deviation of the deaths of components/holes, mean persistence of components/holes, median persistence of components/holes, standard deviation of the persistences of components/holes, minimum birth of components/holes, maximum birth of components/holes, minimum death of components/holes, maximum death of components/holes, range of births of components/holes, range of deaths of components/holes, 1^st^, 5^th^, 25^th^, 50^th^ (median), 75^th^, 95^th^, 99^th^ percentiles of births of components/holes, 1^st^, 5^th^, 25^th^, 50^th^ (median), 75^th^, 95^th^, 99^th^ percentiles of deaths of components/holes. The ratio between the number of holes and the number of components was also computed and added to the list of features, for a total of 57 topological features from each patch.

#### Gabor Filters

Gabor filters are numerical matrices which operate on an image through convolution. They can be described as formed by a spatial frequency and orientation within a two-dimensional Gaussian envelope. For an in-depth explanation of how Gabor filters are used, the reader is referred to [17]. Each Gabor filter is defined uniquely by a direction, a frequency, and the standard deviations in the horizontal and vertical coordinates. Thirty six different Gabor filters of varying directions, frequencies and standard deviations were generated [21]. To compute features using Gabor filters, each image was converted to greyscale and then convolved with the Gabor filters. This yields a filtered greyscale image, from which the mean pixel intensity and variance of pixel intensities were computed. In total 72 Gabor features were computed this way for each patch. The effects that different directions and frequencies for the Gabor filters have on the filtered image are shown in Fig. 5.

**Fig. 5.**
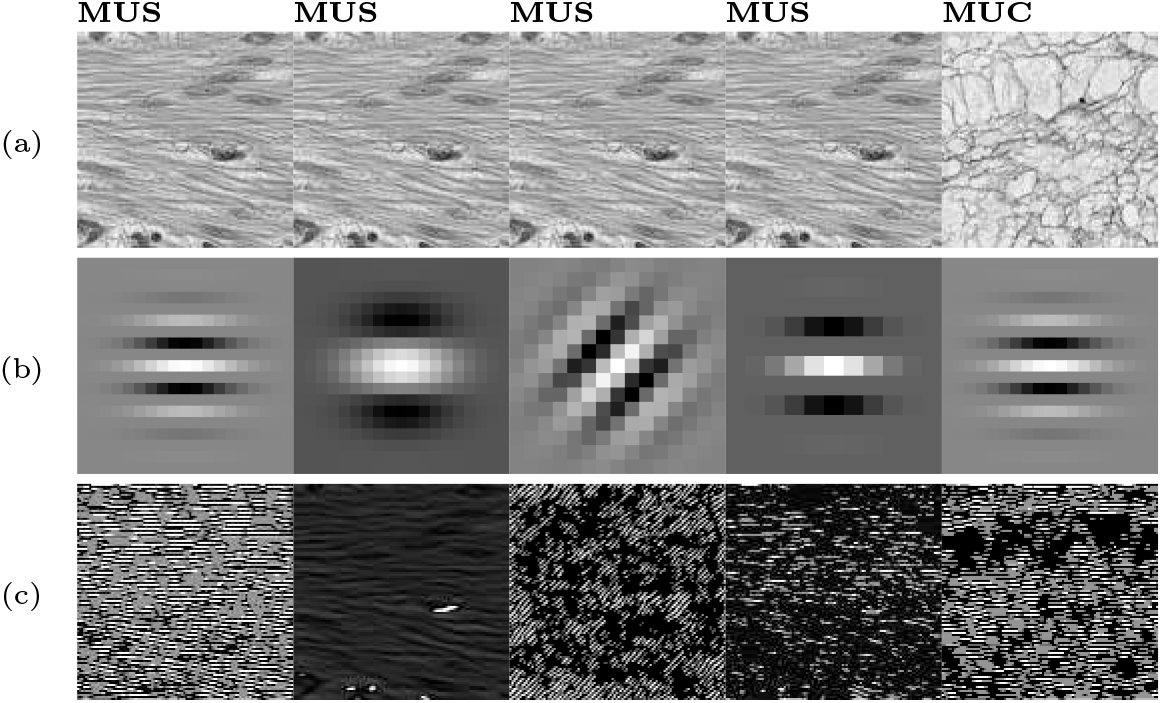
Illustration of extraction of textural characteristics with Gabor filters. (a) Sample patches of the MUS and MUC classes converted to greyscales. (b) Gabor filters with different frequencies and orientations. (c) Filtered results. It should be noticed how certain features are detected by one filter and not the others (e.g., in the first two columns).

The concatenation of the HSV, Topological, and Gabor features, yielded a total of 177 hand-crafted features computed for each patch.

#### Random Forest (RF)

Random Forest (RF) classifiers are a type of classical ML model introduced in 2001 by L. Breiman [4]. They are based on tree classifiers [5] and a technique called “bootstrapping”. By randomly sampling with replacement, many subsets of the training data are created, then each bootstrapped set is used to create a single decision tree, which splits the feature space according to decision boundaries. The process is repeated many times, to create many decision trees, which then “vote” on which label should be assigned to an incoming sample. An in-depth overview can be found in [1].

RF classifiers were trained on the subset of 20,000 samples from the 100K-set and tested on the 7,180 samples of the 7K-set. Then, subsets of up to two classes were chosen to be cleaned according to the process outlined below. The resulting set of samples was then used to train a new RF classifier and tested on the 7K-set. The model was fit using 100 estimators (decision trees) and a maximum depth of 100 nodes for each estimator. The criterion used to build the decision trees was the Gini index. The *out-of-bag* (OOB) score is an accuracy score that is computed by classifying the samples which are not included in the bootstrapped sets for each tree [3]. Both the OOB score and test accuracy were recorded.

#### Residual Network

A Residual Network (ResNet) is a type of convolutional neural network architecture introduced by He et al. [8]. The ResNet-18 architecture offers relatively low computational complexity while maintaining strong performance on image classification tasks. The model was trained on the 100K-set and evaluated on the 7K-set, following a similar protocol as in the RF experiments. Similarly to the Random Forest experiments, the ResNet-18 was trained on the original dataset (without cleaning), and on versions of the dataset that were cleaned according to the process outlined below. However, due to time-constraints and the complexity of training a CNN, only a small subset of classes were chosen to be cleaned. Based on the RF results, the class that yielded the greatest improvement in OOB score was selected for cleaning. Additionally, a visual inspection of the dataset (Fig. 1) suggested that the MUS class exhibited a higher degree of inconsistencies compared to other classes, and was also selected. This selection strategy enables an assessment of whether class-specific improvements observed in classical ML models extend to DL models, as well as whether visually identified inconsistencies correspond to measurable improvements in performance.

The model was trained using standard cross-entropy loss and optimised using the Adam optimiser [13]. Moreover, it is well known that training time and sample size have a large effect on the accuracy and generalisation ability of DL models. So, for these experiments, different sample sizes (5, 000, 10, 000, 20, 000, 50, 000, 100, 000) were tested by splitting the entire training set into as many non-overlapping folds as could be made for each size (e.g. 20 folds for a sample size of 5, 000). The model was trained on each fold and the training and testing accuracies after 5, 10, 15, and 20 epochs were recorded.

#### Cleaning

To test whether cleaning outlier patches from a given class had an effect on ML and DL models, “clean” versions of the 100K-set were created. It must be noted that the RF was trained on a subset of 20,000 images of the 100K-set, while the ResNet-18 was trained on subsets (folds) of different sizes. Each of these subsets (taken as the training set), classes were selected to be cleaned: all classes and all combinations of two classes, for the RF; the highest performing class from the RF and MUS, for the ResNet-18.

For each selected class, the mean and standard deviation of each feature were computed. Any training sample with a feature value over 2.5 standard deviations away from the mean of the given class was considered an outlier and removed. This process yielded a version of the 100K-set that was smaller than the original (since outliers were removed). The particular size of the dataset depended on the classes that were cleaned.

## 3 Results

### Random Forest Experiments

The results of the classification of the datasets with Random Forests are shown in Table 1. The table reports the out-of-bag (OOB) score and test accuracy for models trained on the original dataset after selectively removing outlier samples from one or two classes.

**Table 1.** OOB Scores and test accuracies obtained by each Random Forest model, trained on datasets with and without cleaning. In **bold** are all scores greater than the model trained on the original dataset. <u>Underlined</u> is the highest score.

| Cleaned Classes | Training Samples | OOB Score | Test Accuracy |
| --- | --- | --- | --- |
| None | 20000 | 0.940 | 0.746 |
| STR | 18896 | **<u>0.951</u>** | **0.750** |
| NORM | 19131 | **0.949** | 0.725 |
| TUM | 18546 | **0.946** | 0.708 |
| MUC | 19090 | **0.945** | 0.743 |
| DEB | 18964 | **0.944** | **0.778** |
| MUS | 18602 | **0.944** | 0.738 |
| ADI | 19124 | **0.941** | 0.689 |
| LYM | 18818 | 0.939 | 0.697 |
| BACK | 18734 | 0.937 | 0.685 |
| NORM, STR | 17884 | **0.956** | 0.711 |
| STR, TUM | 17311 | **0.955** | 0.729 |
| DEB, NORM | 17905 | **0.951** | **0.755** |
| DEB, STR | 17833 | **0.950** | **0.790** |
| DEB, TUM | 17508 | **0.950** | **0.777** |
| MUC, STR | 17893 | **0.950** | **0.751** |
| MUS, STR | 17552 | **0.950** | 0.734 |
| NORM, TUM | 17579 | **0.950** | 0.712 |
| LYM, STR | 17712 | **0.950** | 0.707 |
| ADI, STR | 17684 | **0.950** | 0.641 |
| MUS, NORM | 17602 | **0.949** | 0.710 |
| MUS, TUM | 17024 | **0.949** | 0.697 |
| MUC, NORM | 18261 | **0.948** | 0.724 |
| ADI, NORM | 18162 | **0.948** | 0.649 |
| DEB, MUS | 17315 | **0.947** | **0.776** |
| LYM, NORM | 17810 | **0.947** | 0.711 |
| MUC, TUM | 17586 | **0.946** | 0.696 |
| DEB, MUC | 17743 | **0.945** | 0.772 |
| DEB, LYM | 17794 | **0.944** | **0.758** |
| MUC, MUS | 17557 | **0.944** | 0.721 |
| ADI, DEB | 17549 | **0.944** | 0.687 |
| LYM, TUM | 17201 | **0.942** | 0.687 |
| ADI, MUS | 17217 | **0.942** | 0.680 |
| ADI, TUM | 17313 | **0.942** | 0.616 |
| LYM, MUS | 17351 | **0.941** | 0.705 |
| LYM, MUC | 18075 | **0.941** | 0.704 |
| ADI, MUC | 17926 | **0.941** | 0.690 |
| BACK, STR | 17576 | 0.940 | 0.706 |
| BACK, MUC | 17536 | 0.939 | 0.681 |
| BACK, NORM | 17886 | 0.938 | 0.715 |
| BACK, MUS | 17038 | 0.938 | 0.694 |
| ADI, LYM | 17931 | 0.938 | 0.620 |
| BACK, LYM | 17769 | 0.937 | 0.691 |
| BACK, TUM | 17657 | 0.937 | 0.673 |
| BACK, DEB | 17422 | 0.936 | 0.744 |
| ADI, BACK | 17440 | 0.936 | 0.613 |

In the cases where only one class was cleaned, moderate variations in performance were observed. The highest OOB score (0.951) was obtained when cleaning the STR class, while the highest test accuracy (0.778) was achieved when cleaning the DEB class. Cleaning a class increased the OOB Score in seven out of nine cases. However, cleaning only increased test accuracy in two out of the nine classes. When two classes were cleaned, 27 out of 36 cases improved OOB Score but only six cases improved test accuracy. The highest increase in test accuracy, was of 4.45%. In general, cleaning classes such as STR, DEB, MUC, and MUS resulted in higher test accuracies compared to cleaning ADI, LYM, or BACK, which consistently yielded lower performance. The number of training samples varied depending on the class cleaned, this reflects in the number of detected outliers per class.

### ResNet-18 Experiments

The classes selected for cleaning in the ResNet-18 experiments were STR and MUS. STR was chosen because cleaning it yielded the highest improvement in the OOB Score for the Random Forest model. Building on the previous work in [6], the class MUS was also selected to be cleaned because a visual assessment of the patches revealed this class was the one with highest inconsistency (Fig. 1).

The results of the classification of the datasets with ResNet-18 are shown in Fig. 6 as line plots with shaded regions representing the ranges covered by all the folds. The training accuracies across all sample sizes and training times remained similar whether classes were cleaned or not. The behaviour with respect to the training epochs was consistent with normal behaviour: higher number of epochs result in a higher accuracy. Very high training accuracies were achieved even for small training sets of 5,000 samples. This indicates that the network was capable of learning the characteristics of the training data effectively. In contrast, test accuracy was lower and more variable, but higher accuracies were achieved as the number of training samples grew. After 20,000 samples, however, the improvement in test accuracies was less apparent.

**Fig. 6.**
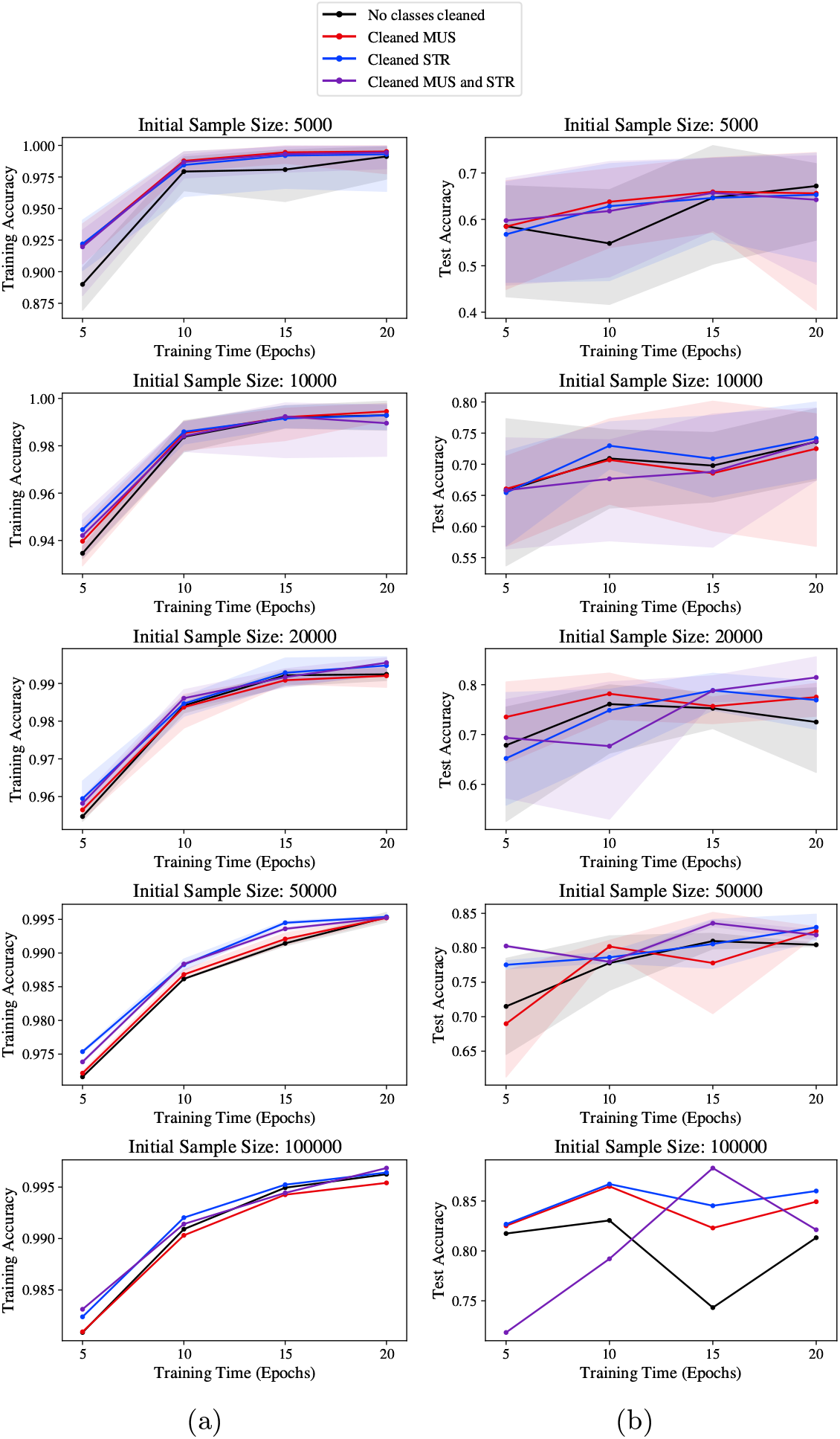
Line graphs show the accuracies obtained for each size of initial training set with and without cleaning. The shaded regions show the ranges obtained by all the folds of the given sample size. Black: models trained on datasets without cleaning. Red: models trained on datasets after cleaning MUS class. Blue: models trained on datasets after cleaning STR class. Purple: models trained on datasets after cleaning MUS and STR. (a) Training accuracy. (b) Test accuracy.

## 4 Discussion

Random Forests rely on splitting the feature space to make classification possible. It is therefore unsurprising that they would be more susceptible to outlier removal than CNNs. This was observed in the experiments (Table 1). However, the experiments also indicated that the effect of outlier removal is not consistent across classes; certain classes improve performance at a much higher rate than others. This was to be expected, as not all classes were found to be overlapping in feature space [6]. However, considerable improvements in the test accuracy were possible when directed at certain classes. For example, cleaning the DEB class only, allowed for an improvement of 3.2 percent (Table 1). The test accuracy was improved by 4.4 percent when both DEB and STR were cleaned. This suggested that the traditional ML model that was selected was able to separate the classes better when certain outliers were discarded. MUS appeared visually to exhibit some inconsistencies in labelling and normalisation (Fig. 1), however, no improvement was exhibited in the test accuracy from the Random Forest, when this class was cleaned.

On the other hand, CNNs learn deep features on the entire image, as the images will include the texture, colour and topological characteristics, in addition to other features that were not specifically extracted, e.g., shapes or patterns. This makes the DL approach robust to noise. This was observed in the experiments as the effect of cleaning classes was much less pronounced on the CNN model. As mentioned previously, the effect on training accuracy was indistinguishable, as the training graphs in Fig. 6 (a) trace very similar paths. The variability among the test accuracies (Fig. 6 (b)) was more evident, but the shapes and scores achieved were not distinct enough to conclude that there is a clear advantage of cleaning the dataset when training neural networks. Additionally, even though there was noticeable improvement when STR was cleaned, this improvement did not carry over to the ResNet-18. Similarly, even though MUS initially appears to be inconsistently labelled and normalised, the experiments did not indicate that this would have a meaningful effect on the generalisation abilities of CNNs.

To refer back to the research questions outlined in Section 1: *to what extent do the outliers, and differences in class separability affect the performance of ML models? In particular, does the presence of samples which may be atypical due to issues such as mislabelling or poor normalisation in certain classes, affect the generalisation of ML models to other datasets?* The results offer the following answers.

1. Outliers and class-specific inconsistencies showed a measurable impact on model performance. However, this effect was mostly notable in traditional ML models such as a Random Forests, where the removal of outliers from specific classes (namely, STR and DEB) yielded noticeable improvements in test accuracy.
2. The impact on the classifications with DL models was less pronounced compared with ML models. Variations in test accuracies were observed but the overall behaviour of the ResNet-18 model remained largely unchanged between the cleaned and the original datasets. This suggests that CNNs are more robust to the presence of outliers.
3. However, with DL, a plateau was observed in test accuracy for larger training sets (Fig. 6 (b)). The plateau suggests that simply increasing the size of a dataset does not fully address the limitations that arise from the quality of the images.
4. As a whole, these findings demonstrate that while data cleaning can be beneficial, particularly for classical models, its impact on DL models may be more subtle and dependent on the nature and extent of the underlying data inconsistencies.

### Limitations and Future Work

It must be recognised that there exist some limitations of the approach that was taken in this study. First, for the experiments with ResNet-18, only a small number of classes was selected to be cleaned, it remains unknown if the selection of another set of classes could have had a bigger impact on the ability of generalisation of the model. Additionally, the cleaning process was based on three types of features, another process of cleaning the dataset may lead to a better generalisation performance from a CNN. Based on these critiques, the conclusions drawn here, should be taken as preliminary results and taken with caution. In future work, we aim to expand the experiments undertaken to incorporate more classes in the cleaning process to evaluate the impact this would have on DL models.

## 5 Conclusion

This study investigated the impact that curation of a dataset can have on the performance of ML and DL models for colorectal cancer histology classification. The results demonstrated that removing outliers from specific classes can lead to measurable improvements in classical ML models, particularly when class inconsistencies are pronounced. In contrast, DL models such as ResNet-18 do not receive as great a benefit, with only marginal changes in performance observes. These findings highlight the importance of data quality for traditional models but also the robustness of DL models.

## Data Availability

All data produced in the present study are available upon reasonable request to the authors.

## Code and Data Availability

Code is available at https://github.com/britod97/Tissue-Classification and data is freely available Zenodo [10].

## Acknowledgements

The authors gratefully acknowledge Prof. Manuel Salto-Tellez, Dr. Tom Lund, Dr. Ferrán Cardoso, Dr. Priya Narayanan, and Dr. Elena Safrygina from the Integrated Pathology Unit, The Institute of Cancer Research (ICR) for their valuable input that contributed to this work.

